# Biological age in UK Biobank: biomarker composition and prediction of mortality, coronary heart disease and hospital admissions

**DOI:** 10.1101/2019.12.12.19014720

**Authors:** Mei Sum Chan, Matthew Arnold, Alison Offer, Imen Hammami, Marion Mafham, Jane Armitage, Rafael Perera, Sarah Parish

**Affiliations:** Nuffield Department of Population Health, University of Oxford, Oxford, United Kingdom; British Heart Foundation Cardiovascular Epidemiology Unit, Department of Public Health and Primary Care, University of Cambridge, Cambridge, United Kingdom; MRC Population Health Research Unit, University of Oxford, Oxford, United Kingdom; Nuffield Department of Primary Health Care Sciences, University of Oxford, Oxford, United Kingdom

## Abstract

**Background:** Age is the strongest risk factor for most chronic diseases, and yet individuals may age at different rates biologically. A biological age formed from biomarkers may be a stronger risk factor than chronological age and understanding what factors contribute to it could provide insight into new opportunities for disease prevention.

**Methods and findings:** Among 480,019 UK Biobank participants aged 40-70 recruited in 2006-2010 and followed up for 6-12 years via linked death registry and secondary care records, a subpopulation of 141,254 (29.4%) non-smoking adults in good health and with no medication use or disease history at baseline were identified. Independent components of 72 biomarkers measured at baseline were characterised by principal component analysis. The Klemera Doubal method (KDM), which derived a weighted sum of biomarker principal components based on the strengths of their linear associations with chronological age, was used to derive sex-specific biological ages in this healthy subpopulation. The proportions of the overall biological and chronological age effects on mortality, coronary heart disease and age-related non-fatal hospital admissions (based on a hospital frailty index) that were explained by biological age were assessed using log-likelihoods of proportional hazards models.

Reduced lung function, reduced kidney function, slower reaction time, lower insulin-like-growth factor 1, lower hand grip strength and higher blood pressure were key contributors to biological age (explaining the highest percentages of its variance) in both men and women, while lower albumin, higher sex hormone-binding globulin and lower muscle mass in men, and higher liver enzymes, blood lipids and HbA1c in women were also important. Across both sexes, a 51-principal component biological age explained 66%, 80% and 63% of the age effects on mortality, coronary heart disease and hospital admissions, respectively. Restricting the biological age to the 12-13 key biomarkers corresponding to the 10 most importantly contributing principal components resulted in little change in these proportions for women, but a reduction to 53%, 63% and 50%, respectively, for men.

**Conclusions:** This study identified that markers of impaired function in a range of organs account for a substantial proportion of the apparent effect of age on disease and hospital admissions. It supports a broader, multi-system approach to research and prevention of diseases of ageing.

## Introduction

Age is the strongest risk factor for most chronic diseases that limit healthy lifespan, yet individuals may age biologically at different rates,^1^ illustrated by differential rates of disease accumulation and frailty onset. Understanding the contributors to biological ageing could lead to opportunities for early prevention of later life disease^2^ and limiting the disease burden associated with growing elderly populations.

‘Biological age’ is often used to refer to a composite risk score estimated from measurements of biomarkers of ageing,^1,3-5^ which may be constructed from different types and numbers of biomarkers.^5-7^ Biological ages constructed from clinical biomarkers widely measured in routine care were found to be more predictive of health outcomes than those constructed using epigenetic ages^2,6^ and telomere length.^6^ The most important contributors to biological age have varied between studies, in part due to differences in the set of biomarkers available.^8,9^

The main methods of estimating biological ages using clinical biomarkers are multiple linear regression, principal component analysis (PCA) and the Klemera Doubal method (KDM), which derives a weighted sum of biomarkers based on the strengths of their associations with chronological age^10^ and is favoured by comparison studies.^9,11,12^ KDM allows comparison of the predictive value of its biological age with that of chronolological age for health outcomes, when chronological age has not been included as a constituent ‘biomarker’. However, KDM biological ages have typically included chronological age^13^ and the predictive values of biological ages have only occasionally been compared with those of chronological age^13^ or mortality-based scores.^2,14^ Multiple linear regression has been used in many studies^5,9,12^ and both multiple linear regression and KDM have been extended to principal components of biomarkers instead of individual biomarkers.^11^

The UK Biobank is a richly phenotyped resource with 0.5 million participants^15^ that provides an unrivalled opportunity to investigate earlier stages of ageing through biological, lifestyle and environmental factors, compared to previous studies of typically 100-10,000 participants with panels of fewer than 30 biomarkers.^5^ A substantial middle aged, apparently healthy subpopulation of the UK Biobank can be identified, to reduce reverse causality from prior health or medication use affecting biomarker levels.

This study aims to: (1) investigate and implement reliable methods of estimating biological age in the UK Biobank, (2) identify the main biomarker determinants of biological age, and (3) investigate the relationship between biological and chronological age in the prediction of health outcomes.

## Methods

### Study population

The UK Biobank recruited 0.5 million participants across the UK aged 40-70 for baseline assessment in 2006-2010. Information on sociodemographic characteristics, self-reported health behaviours and medication was recorded. Linkage to Hospital Episode Statistics (HES) and national death registries provided prior and prospective information on secondary care outcomes and date and cause of death. Over 100 biomarkers and physical attributes were measured using various measurement devices, and blood and urine assays (S1 Appendix 1 and 2).^15^ This study was covered by the general ethical approval for UK Biobank studies from the NHS National Research Ethics Service on 17th June 2011 (Ref 11/NW/0382).

After data cleaning (S1 Figure 1), 480,019 participants, followed up for a median of 8.7 years for death registry records and 8.0 years for HES records were included (S1 Appendix 1). A composite measure of prior health, derived from chronic disease-related medication count, walking speed, number of HES episodes prior to recruitment, smoking status, previous age-related chronic disease and hip or wrist fracture was used to stratify the population into 4 groups: (1) healthy (generally non-smokers with no prior disease or medications), (2) some medications, (3) slightly unhealthy, and (4) poor health (S1 Appendix 1). This study focused on the 141,254 people in the healthy subpopulation, in order to reduce reverse causality, and contrasted results for this subpopulation with those for the poor health subpopulation and the whole population.

### Statistical Analyses

Among 110 available biomarkers, a panel of 72 biomarkers was obtained after excluding biomarkers that: (1) were missing in >30% participants, (2) were measuring the same biological trait as another biomarker, or (3) had poor reproducibility, assessed through intra-individual correlations of <0.1 in the 19,335 individuals with a repeat measurement (S1 Appendix 2 and S1 Table 3). For each sex and prior health subpopulation, trends of each biomarker with chronological age were visually assessed for linearity (S1 Appendix 3A), before using linear methods to estimate biological ages. To represent the biomarkers as linearly uncorrelated principal components, PCA with varimax rotation^11^ was carried out on the 72 biomarkers, which gave 51 principal components with eigenvalues >0.33. These principal components were characterised based on constituent biomarkers with the largest factor loadings (S1 Appendix 3B–D).

Biological ages were estimated using KDM,^10^ separately within each sex and prior health subpopulation (S1 Appendix 3C). A similar method, stepwise linear regression, was considered and the results for both methods were compared in the appendices (S1 Appendix 3C and 4). Biomarker principal components were ranked by their importance, measured by the proportion of variance in the biological ages that each component explained (S1 Appendix 3E).

Three health outcomes were constructed from HES and death records: (1) death from chronic disease (excluding: infectious diseases, pregnancy, congenital malformations and external causes),^16^ (2) fatal and non-fatal coronary heart disease (CHD) and (3) age-related non-fatal hospital admissions (S1 Appendix 2). These hospital admissions are the subset of those types of admissions in a published hospital frailty risk score^17^ that are age-related in the UK Biobank (S1 Table 6). The predictive powers of chronological age and biological ages for each health outcome were assessed using Cox proportional hazards models adjusted for Index of Multiple Deprivation 2010 quintile, smoker status, alcohol consumption and assessment centre, and computing Harrell’s C-indices (measures of statistical discrimination similar to the area under the receiver operating curve). Prediction of CHD and hospital admissions by biological age was compared with a benchmark of prediction by a mortality score similar to those proposed by previous studies^2,14^ and derived from stepwise Cox regression, using unadjusted Cox models (S1 Appendix 3F).

To investigate the relationship of the biological ages to chronological age, the proportion of variation in chronological age described by each biological age was estimated. The proportion of the overall biological and chronological age effect on mortality, CHD and hospital admission risk that was explained by each biological age was also estimated, by comparing the log-likelihoods from these Cox models (S1 Appendix 3G). Calibration of biological ages to chronological age and the risk calibration of biological ages with each health outcome was assessed (S1 Appendix 3H).

The statistical analysis was repeated on biomarkers corresponding to the 10 most important biomarker principal components in the analyses of the panel of 51 biomarker principal components among healthy participants of each sex. The predictive and explanatory power for this reduced biomarker panel was compared to the full panel. Guidelines for Transparent Reporting of a multivariable prediction model for Individual Prognosis Or Diagnosis (TRIPOD)^18^ were followed (S1 Table 14).

## Results

### Study characteristics

Of the 480,019 participants, 141,254 (29.4%) were in the healthy subpopulation (Table 1). During a median follow up period of 8.7 years for mortality and 8.0 years for CHD and hospital admissions (S1 Appendix 1), 1.7% of healthy, and 3.9% of all participants died from chronic diseases; 1.9% of healthy and 4.0% of all participants without CHD at baseline had a first CHD event; 16.0% of healthy and 23.1% of all participants who were not admitted to hospital for age-related reasons prior to baseline had been admitted with diagnoses of these conditions during follow up (S1 Table 7). Sociodemographic patterns and the proportion of participants healthy at baseline were similar between sexes.

**Table 1:** Participant characteristics at baseline assessment in 2006-2010, for the Healthy and Poor health subpopulations and the whole UK Biobank population

|  | Healthy subpopulation |  |  | Poor health subpopulation |  |  | Whole population |  |  |
| --- | --- | --- | --- | --- | --- | --- | --- | --- | --- |
|  | Persons | Men | Women | Persons | Men | Women | Persons | Men | Women |
| **Participants (n)** | 141,254 | 65,869 | 75,385 | 82,835 | 42,277 | 40,558 | 480,019 | 219,248 | 260,771 |
| **Person-years at risk (millions)** | 1.2 | 0.56 | 0.64 | 0.71 | 0.36 | 0.35 | 4.12 | 1.87 | 2.25 |
| **Deaths during follow up (%)** | 1.7 | 2.1 | 1.4 | 9.1 | 11.2 | 7.0 | 3.9 | 5.2 | 2.9 |
| **Median age at baseline (years)** | 56.0 | 55.7 | 56.4 | 62.2 | 62.5 | 61.8 | 58.3 | 58.8 | 58.0 |
| **Age band at baseline in years (%)** |  |  |  |  |  |  |  |  |  |
| 40-44 | 12.1 | 13.9 | 10.5 | 3.9 | 3.9 | 3.9 | 10.2 | 10.4 | 10.1 |
| 45-49 | 15.9 | 16.5 | 15.4 | 6.5 | 6.2 | 6.8 | 13.1 | 12.7 | 13.4 |
| 50-54 | 17.8 | 17.1 | 18.4 | 10.5 | 9.7 | 11.3 | 15.1 | 14.4 | 15.8 |
| 55-59 | 19.4 | 18.6 | 20.1 | 16.8 | 15.7 | 17.9 | 18.1 | 17.5 | 18.6 |
| 60-64 | 21.9 | 21.0 | 22.6 | 30.2 | 30.0 | 30.4 | 24.3 | 24.3 | 24.3 |
| 65-70 | 12.9 | 12.9 | 13.0 | 32.1 | 34.5 | 29.6 | 19.2 | 20.8 | 17.9 |
| **Index of Multiple Deprivation quintile (%)** |  |  |  |  |  |  |  |  |  |
| Q1 (least deprived) | 23.9 | 23.8 | 24.1 | 16.9 | 16.7 | 17.1 | 20.0 | 19.7 | 20.1 |
| Q2 | 22.2 | 22.1 | 22.3 | 18.3 | 18.2 | 18.3 | 20.0 | 19.7 | 20.3 |
| Q3 | 20.9 | 20.7 | 21.1 | 19.3 | 19.2 | 19.4 | 20.0 | 19.8 | 20.2 |
| Q4 | 18.7 | 18.7 | 18.7 | 21.0 | 20.8 | 21.1 | 20.0 | 19.9 | 20.1 |
| Q5 (most deprived) | 14.3 | 14.7 | 13.9 | 24.6 | 25.1 | 24.0 | 20.0 | 20.9 | 19.3 |
| **Smoker status (%)** |  |  |  |  |  |  |  |  |  |
| Current | 0.0 | 0.0 | 0.0 | 11.0 | 12.5 | 9.6 | 10.5 | 12.4 | 8.8 |
| Previous | 33.4 | 35.9 | 31.2 | 41.6 | 47.1 | 35.9 | 34.5 | 38.3 | 31.3 |
| Never | 66.6 | 64.1 | 68.8 | 46.6 | 39.7 | 53.7 | 54.5 | 48.8 | 59.4 |
| No answer/missing | 0.0 | 0.0 | 0.0 | 0.8 | 0.8 | 0.7 | 0.5 | 0.5 | 0.5 |
| **Alcohol consumption frequency (%)** |  |  |  |  |  |  |  |  |  |
| Never | 5.5 | 4.5 | 6.5 | 11.5 | 8.8 | 14.2 | 8.0 | 6.3 | 9.5 |
| Special occasions only | 8.8 | 5.6 | 11.6 | 14.1 | 9.3 | 19.1 | 11.5 | 7.3 | 15.0 |
| One to three times a month | 10.5 | 8.6 | 12.3 | 10.8 | 9.1 | 12.6 | 11.1 | 8.9 | 13.0 |
| Once or twice a week | 27.3 | 27.2 | 27.4 | 24.5 | 25.5 | 23.5 | 25.8 | 25.9 | 25.7 |
| Three or four times a week | 26.8 | 29.5 | 24.5 | 19.8 | 23.1 | 16.4 | 23.1 | 26.1 | 20.5 |
| Daily or almost daily | 21.0 | 24.7 | 17.7 | 19.0 | 23.8 | 14.0 | 20.3 | 25.3 | 16.1 |
| No answer/missing | 0.0 | 0.0 | 0.0 | 0.3 | 0.3 | 0.3 | 0.2 | 0.2 | 0.2 |
| **Body mass index in kg/m<sup>2</sup> (%)** |  |  |  |  |  |  |  |  |  |
| <18.5 | 0.6 | 0.2 | 0.9 | 0.5 | 0.3 | 0.7 | 0.5 | 0.2 | 0.8 |
| 18.5-24.9 | 41.7 | 32.6 | 49.6 | 24.5 | 19.6 | 29.7 | 32.8 | 25.2 | 39.2 |
| 25-29.9 | 43.1 | 51.6 | 35.8 | 41.5 | 46.2 | 36.6 | 42.4 | 49.2 | 36.6 |
| 30-39.9 | 14.1 | 15.3 | 13.0 | 30.1 | 31.4 | 28.7 | 22.4 | 24.0 | 21.0 |
| 40+ | 0.5 | 0.3 | 0.7 | 3.4 | 2.5 | 4.3 | 1.9 | 1.3 | 2.4 |

### Biomarker characteristics

The relationships of most candidate biomarkers to chronological age were broadly linear or flat (S1 Figure 3 and Figure 1). Several biomarkers displaying non-linear trends and differences by sex or by prior health are highlighted in Figure 1, which shows standardised biomarker levels for comparability between biomarkers. Lung function biomarkers, systolic blood pressure, cystatin C and reaction time had the strongest linear relationships with age, while diastolic blood pressure, body mass index (BMI) and low density lipoprotein cholesterol (LDL-C) had clear inverse U-shaped relationships with age that were attenuated in the healthy subpopulation. Diastolic blood pressure peaked or plateaued at different ages and BMI displayed different trends in the healthy and poor health subpopulation. Biomarkers that displayed substantially different trends between sexes were heel bone density, LDL-C, calcium, alkaline phosphatase and phosphate (S1 Figure 3).

**Figure 1:**
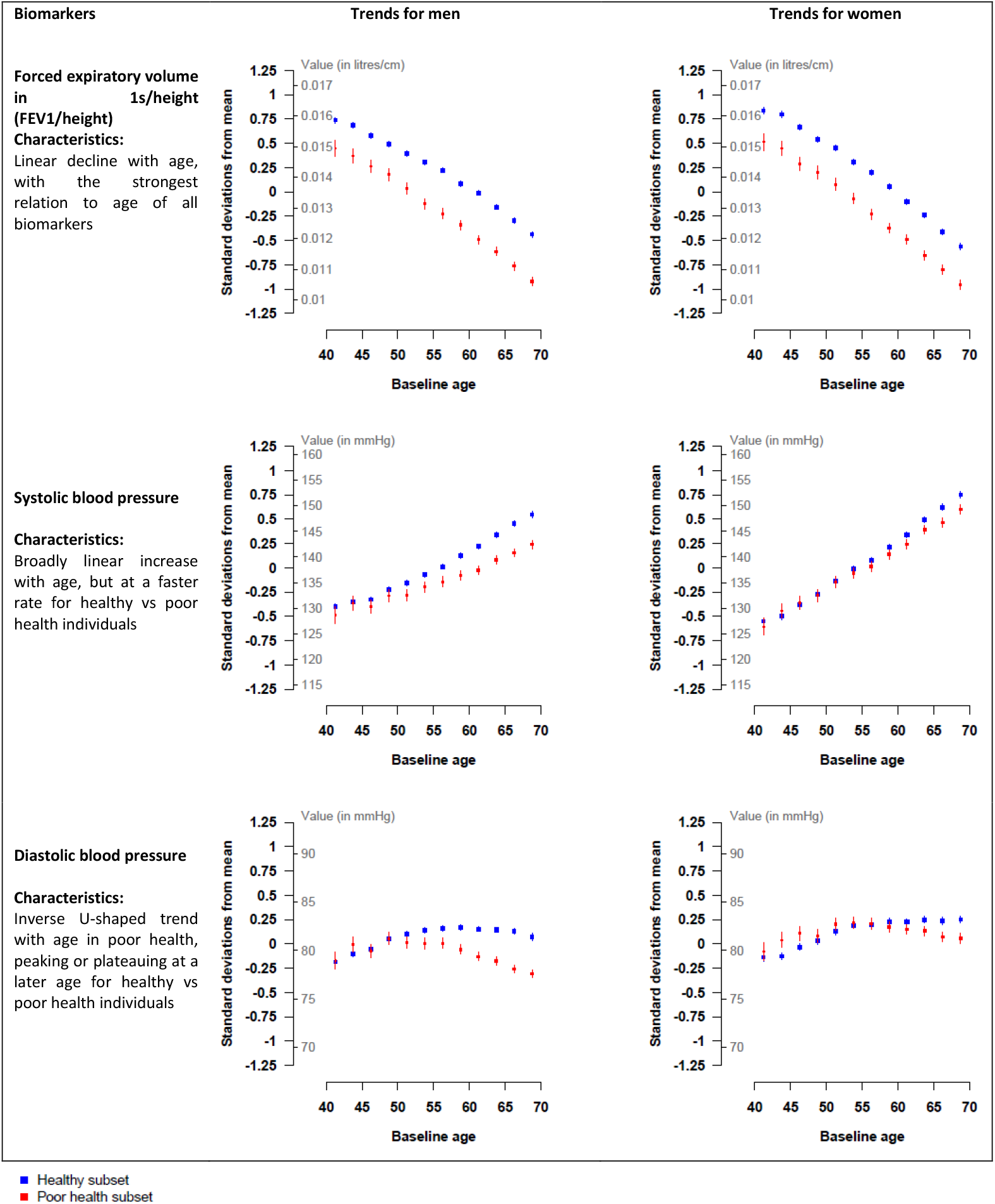
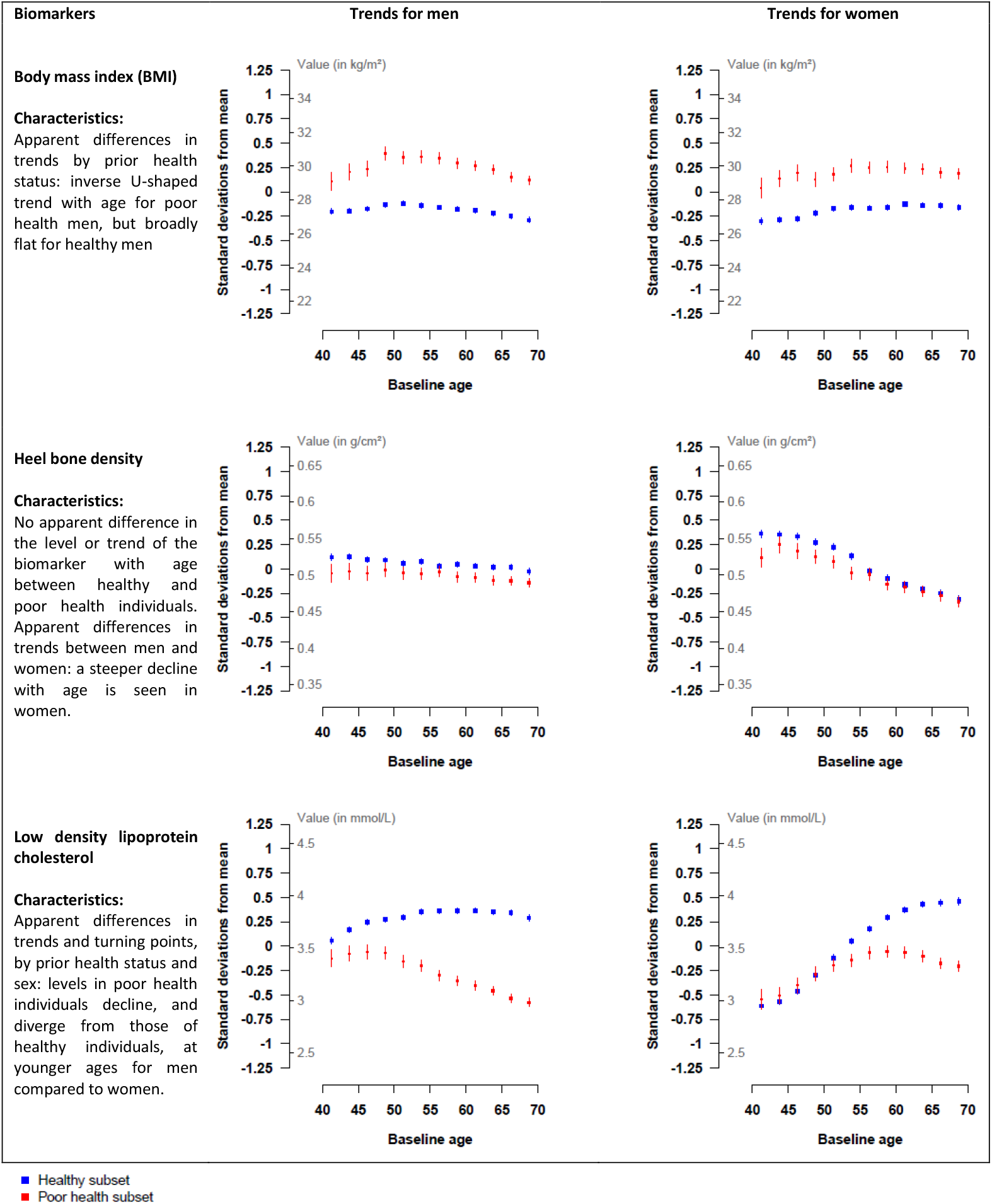
Biomarker-age trends (means and standard errors for each 2.5-year age group) for the healthy and poor health subpopulations and descriptions of their characteristics, for six selected biomarkers and by sex

Many biomarker principal components had a single biomarker strongly loaded onto them and were easily characterised. Multiple biomarkers were strongly loaded onto the adiposity, lung function, blood pressure and blood lipid principal components (S1 Figure 5). The coefficients of the biomarker principal components in the biological ages for the healthy subpopulation are listed in S1 Table 9. The estimated biological ages appeared stable in 10-fold cross validation as their prediction errors were small (S1 Appendix 4 and S1 Figure 6).

### Predictive power and calibration of biological ages

The KDM biological ages (based on 51 principal components) were well calibrated as they matched healthy participants’ chronological ages on average (S1 Figure 7). The KDM ages were more predictive of CHD and hospital admissions than the benchmark mortality score (approximate increases in C-indices for CHD/hospital admissions: 0.135/0.111 in men, 0.109/0.068 in women), and the mortality score performed only slightly better than chance (C-indices ∼0.5; S1 Table 12). KDM ages alone were not statistically significantly better than chronological age and did not statistically significantly supplement chronological age alone in predicting the health outcomes in the healthy subpopulation (increase in C-indices for mortality/CHD/hospital admissions: 0.007/0.006/0.002 in men, 0.002/0.023/0.001 in women; S1 Table 13). However, when estimated in unhealthier subpopulations, they supplemented chronological age alone in the prediction of mortality and CHD in these subpopulations (for the whole population, increase in C-indices: 0.031/0.015 in men, 0.014/0.028 in women; S1 Table 13).

The stepwise regression age was poorly calibrated as it was distributed across a narrower age range than chronological age on average (S1 Figure 7), and was not considered in further analyses. The prediction results and a risk calibration assessment are described in detail in S1 Appendix 4.

### Biomarker importance in biological ages

In the KDM ages, reduced lung function featured most strongly in the healthy subpopulation (Figure 2), describing 12.4% (men) and 10.3% (women) of the variation in biological age (S1 Table 10). Higher cystatin C, slower reaction time, lower insulin-like growth factor-1 (IGF-1), lower hand grip strength, higher and higher blood pressure also featured strongly for both sexes; while lower albumin, higher sex hormone-binding globulin and lower muscle mass biomarkers featured strongly for men; and higher levels of alkaline phosphatase, LDL-C and apolipoprotein B and HbA1c for women. Multiple body systems were represented by these biomarkers: respiratory, renal, cardiovascular, musculoskeletal, endocrine, metabolic and immune, liver and nervous systems (S1 Table 5). When the analysis was restricted to the top 10 biomarker components corresponding to 13 biomarkers for men and 12 for women, forced expiratory volume in 1s/height, forced vital capacity/height, reaction time, IGF-1, cystatin C, hand grip strength/height, systolic and diastolic blood pressure in both sexes; albumin, sex hormone-binding globulin, fat-free mass, standing height and sitting height in men; and LDL-C, alkaline phosphatase, HbA1c and urea in women featured most strongly.

**Figure 2:**
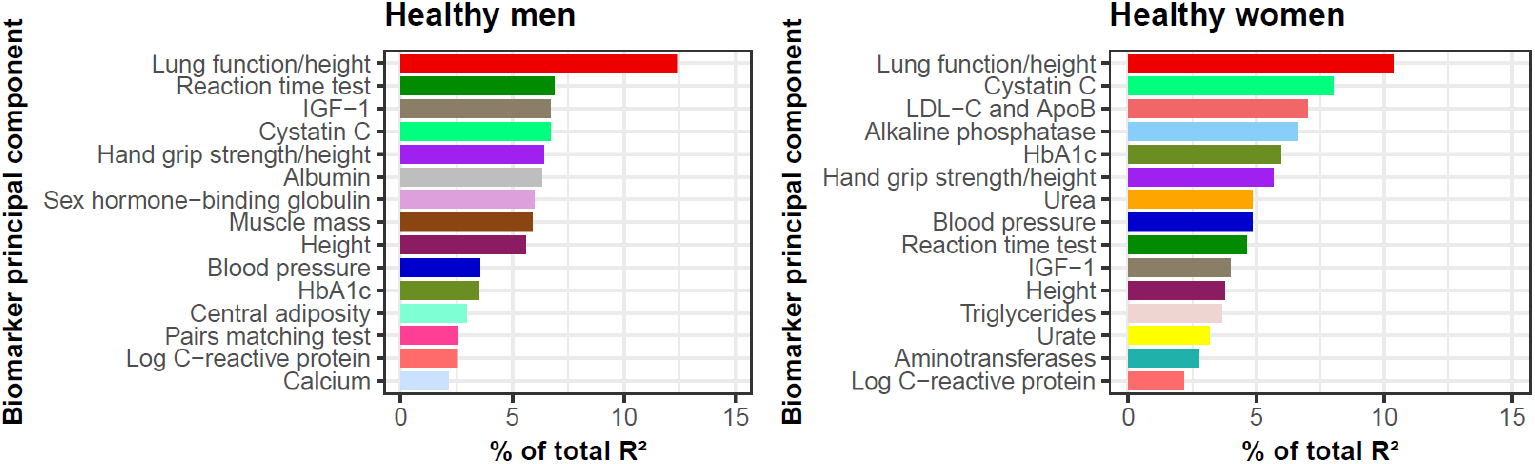
Importances of the top 15 biomarker principal components in the Klemera Doubal (KDM) biological ages for healthy men (left) and women (right)

### Relationship between biological and chronological age

The KDM ages described 44.0% and 51.3% of the variation in chronological age for healthy men and women respectively. More importantly, with respect to the prediction of mortality, CHD and hospital admissions, and averaged across sexes, the KDM ages described 66%, 80% and 63% of the overall biological and chronological age effect respectively (Figure 3A and S1 Table 11). The proportion described by the KDM age is attributed to each constituent biomarker, broadly in the proportions of biomarker importances (Figure 2). Constructing the KDM age from the reduced panel of 12-13 biomarkers decreased the proportion explained by biomarkers to 53%, 63% and 50% for each respective outcome in men, but made little difference for women (Figure 3).

**Figure 3:**
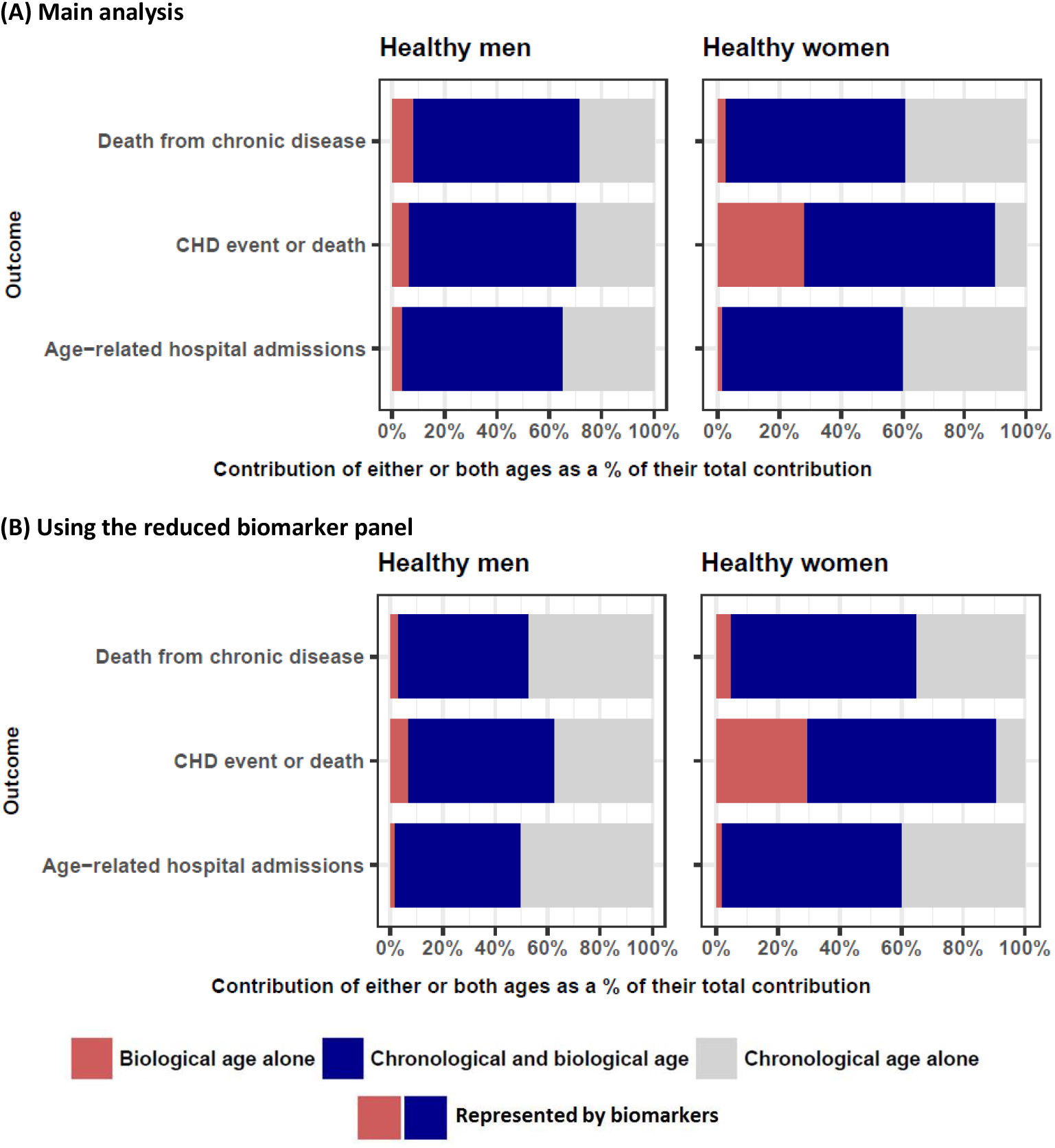
Relative contribution of Klemera Doubal (KDM) biological ages and chronological age in explaining each health outcome, in the (A) main analysis (top) and when (B) using the reduced biomarker panel (bottom), for healthy men (left) and women (right)

## Discussion

This study found that biological ages consisting of markers of impaired function in a range of organs accounted for a substantial proportion of the apparent effect of chronological age on disease and hospital admissions.

### Key biomarker determinants of biological ages

Lung, kidney, cognitive and liver function, IGF-1, hand grip strength and blood pressure were key contributors to biological ages for both sexes, while sex hormone-binding globulin and muscle mass in men, and cardiovascular function and HbA1c in women were also important.

The top-ranking biomarkers in this UK population generally matched those in a Singaporean study^9^ and slight variations by sex were seen in both populations. However, these lung and renal function biomarkers were not investigated in the study of Canadian, South Korean and Eastern European biological ages, which instead found that the top-ranking blood-based biomarkers varied by population and sex.^8^ Studies of ageing biomarkers also found that lung and renal biomarkers were top-ranking determinants of functional decline^19^ and variation in age-related traits.^20^ The present study provides additional detail on the relative importance on ageing of biomarkers within body system groups, such as cystatin C over other renal biomarkers (creatinine and creatinine-based eGFR),^21^ as previous studies each assessed only one of these biomarkers.^8,9,19,20^

Several key biomarkers in this study (blood pressure, blood lipids, height and lung function) have each been shown to be associated observationally, and in some cases causally, in randomised trials and Mendelian randomisation studies, with a range of age-related diseases (Table 2). Associations for other key determinants such as cystatin C and hand grip strength have been less extensively researched, and available studies have focused on mortality and cardiovascular outcomes.^21-25^ Blood pressure did not feature as strongly as the aforementioned biomarkers in our study, despite being well-established as a modifiable and causal risk factor of cardiovascular disease.^26^ Other cardiovascular biomarkers did not feature strongly for men, whereas for women, LDL-C and apolipoprotein B, causally linked to atherosclerotic cardiovascular disease,^27^ were also important (Figure 2), and may have contributed to better prediction of CHD in women than in men (S1 Tables 12 and 13). Cohort effects in this population are difficult to disentangle, and may influence trends in body size. Hence, height may be acting as a proxy for cohort effects.

**Table 2:** Examples of associations in published studies between the top 10 biomarker principal components of biological age identified in the present study and adverse health outcomes

| Key biomarker principal component in the present study | Evidence from meta-analyses of randomised trials | Evidence from Mendelian randomisation | Evidence from prospective studies |
| --- | --- | --- | --- |
| Lung function/height | - | Respiratory and autoimmune diseases<sup>28</sup> | All-cause, circulatory disease, respiratory and cancer mortality<sup>29</sup> |
| Cystatin C | - | (Not significantly associated with cardiovascular disease<sup>23</sup>) | Cardiovascular disease, mortality<sup>21,22</sup> and end-stage renal disease<sup>22</sup> |
| Reaction time test | - | - | Mortality<sup>30</sup> |
| IGF-1 | Fracture risk<sup>31</sup> | (Not significantly associated with Alzheimer's disease<sup>32</sup>) | Mortality and heart failure<sup>33</sup> and cognitive function<sup>34</sup> |
| Hand grip strength/height | - | Cardiovascular disease and mortality<sup>24</sup> | Mortality, cardiovascular disease, respiratory disease, cancer<sup>25</sup> |
| Blood pressure | Mortality and cardiovascular disease<sup>26</sup> | Type 2 diabetes,<sup>35</sup> Alzheimer's disease<sup>36</sup> | Vascular mortality (associations weaken with age)<sup>37</sup> |
| Albumin (men only) | - | - | Coronary heart disease<sup>38</sup> |
| Sex hormone-binding globulin (men only) | - | Type 2 diabetes<sup>39</sup> | Type 2 diabetes<sup>40</sup> |
| Muscle mass (men only) | - | - | Cancer mortality,<sup>41</sup> physical disability<sup>42</sup> |
| Height (men only) | - | Cardiovascular disease, hip fracture, intervertebral disc disorder, vasculitis, gastro-oesophageal reflux disease and cancer<sup>43</sup> | Mortality from cardiovascular disease, liver disease, COPD, stomach and oral cancers, mental disorders<sup>44</sup> |
| LDL-C and ApoB (women only) | Major vascular disease, vascular and all-cause mortality<sup>45</sup> | Coronary heart disease<sup>46</sup> | Cardiovascular disease<sup>47</sup> |
| Alkaline phosphatase (women only) | - | Type 2 diabetes<sup>48</sup> (Associations with IHD and T2D not robust after allowing for pleiotropy<sup>49</sup>) | Osteosarcoma<sup>50</sup> and cardiovascular disease<sup>51</sup> |
| HbA1c (women only) | (Not significantly associated with cardiovascular disease<sup>52</sup>) | Coronary artery disease<sup>53</sup> | Mortality, cardiovascular disease, cancer, diabetes<sup>54</sup> |
| Urea (women only) | - | - | Coronary heart disease<sup>55</sup> |

### Relationships of biomarkers to age

Lung function biomarkers, systolic blood pressure, cystatin C and reaction time had the strongest linear relationships with age regardless of prior health, and therefore contributed substantially to variation in the KDM biological ages (Figure 2). The differences in trends for diastolic blood pressure, BMI and LDL-C by prior health were suggestive of reverse causal effects of disease or medication use on biomarker levels. BMI featured less strongly in the biological ages (the general adiposity component was 28^th^ most important for men and 26^th^ for women) potentially due to reverse causality,^56^ but has been causally linked to 30 diseases.^57^ Therefore, BMI may be a modifiable risk factor that affects biological age.

### Prediction in healthy versus unhealthy individuals

Knowledge of biological age is potentially more useful in apparently healthier individuals because in unhealthy individuals, disease and hospital admissions are diagnostic indicators of ageing;^58^ furthermore knowledge of risk of non-fatal outcomes is likely to provide a longer window for intervention and prevention than knowledge of mortality risk. Biological ages were better than the benchmark mortality score in predicting CHD and hospital admissions (S1 Table 12). The predictive value of a biological age varied by health status and was greater in unhealthy individuals (S1 Tables 12 and 13), likely reflecting the contribution of diagnostic indicators of ageing. Therefore, it is important to take into account the health and age profile of the population when comparing different studies.

The KDM permitted investigation of the relationship between biological and chronological ages with respect to predicting health outcomes, and automatically calibrated biological ages to chronological age. By contrast, the stepwise regression method resulted in a poorly-calibrated biological age in the UK Biobank despite its frequent use in biological ageing studies^5,9,12^ (where its risk calibration was not investigated). Comparison of an individual’s biological age with their chronological age could provide a valuable means of communicating modifiable health risks, alongside their detailed biomarker profile.^59^ A prognostic biological age could also augment a national prevention programme promoting clinical biomarker screening in a middle-aged population,^60^ after causal factors underlying its constituents have been established. The most important biomarkers in biological age were measured via blood biochemistry measurements, spirometry and physical measurements, which can currently be administered quickly and simultaneously in clinical settings. For women, it may be suitable to measure just 12 key biomarkers across 7 body systems to assess biological ageing, as relatively little explanatory and predictive value was compromised. Despite the successful use of clinical risk prediction tools, ‘heart age’ and ‘lung age’ in clinical care,^61^ there is little evidence as yet of implementation of an overall biological age, and proposals to use it in drug development^8,16,62^ and clinical care^2,5,62^ may be longer term uses.

### Strengths and limitations of this study

The estimation methods used assumed that biomarkers with the strongest linear relation to chronological age contribute most to biological age, but these biomarkers are not necessarily strongly linked to health outcomes. Analysis of key determinants of biological age were limited by the range of biomarkers available. Not all biomarker trends in the UK Biobank were linear (S1 Figure 3), but a previous study has shown that incorporating non-linearity was computationally complex and only slightly improved the accuracy of estimated biological age components.^63^ However, the epidemiological reliability of the present analyses was increased through stratification by prior health, use of biomarker principal components, cross validation and adherence to clinical risk prediction reporting guidelines (S1 Table 14).^18^

## Conclusions

A biological age consisting of clinical biomarkers reflecting functionality of a range of organs accounted for a substantial proportion of the effect of age on disease and hospital admissions in the UK Biobank. An overall biological age has potential to be used and evaluated as a broader-based approach to risk identification and prevention than individual biomarkers. Of the most important biomarkers contributing to the derived biological age, cardiometabolic biomarkers have well-studied causal associations with mortality and cardiovascular disease, but further research is needed to identify modifiable causal factors underlying all components, for a range of age-related diseases.

## Data Availability

The underlying data is open access through application to the UK Biobank, and materials and methods will be made freely available through the UK Biobank as part of this project.

## Acknowledgements

This research used the UK Biobank resource (application number 8835). We thank the participants of UK Biobank for their contribution to the resource. The work of MA was supported by core funding from: the UK Medical Research Council (MR/L003120/1), the British Heart Foundation (RG/13/13/30194; RG/18/13/33946) and the National Institute for Health Research [Cambridge Biomedical Research Centre at the Cambridge University Hospitals NHS Foundation Trust]. The views expressed are those of the authors and not necessarily those of the NHS, the NIHR or the Department of Health and Social Care.

